# TRENDS IN ACCESS TO WATER AND SANITATION IN MALAWI: PROGRESS AND INEQUALITIES (1992-2017)

**DOI:** 10.1101/2020.03.30.20047613

**Authors:** Alexandra Cassivi, Elizabeth Tilley, E.O.D. Waygood, Caetano Dorea

**Author notes:** **Corresponding author:** Alexandra Cassivi - University of Victoria, Department of Civil Engineering, Engineering and Computer Science (ECS) 304, PO Box 1700 STN CSC, Victoria BC V8W 2Y2, Canada –.

## Abstract

Billions of people globally gained access to improved drinking water sources and sanitation in the last decades, following effort towards the Millennium Development Goals. Global progress remains a general indicator as it is unclear if access is equitable across groups of the population. Agenda 2030 calling for “leaving no one behind”, there is a need to focus on the variations of access in different groups of the population, especially in the context of least developed countries including Malawi. We analyzed data from Demographic Health Survey (DHS) and Multiple Indicator Cluster Survey (MICS) to describe emerging trends on progress and inequalities in water supply and sanitation services over a 25-year period (1992 - 2017) and to identify the most vulnerable population in Malawi. Data were disaggregated with geographic and socio-economic characteristics including regions, urban and rural areas, wealth and education level. Analysis of available data revealed progress in access to water and sanitation among all groups of the population. The largest progress is generally observed in the groups that were further behind at the baseline year, which likely reflects good targeting in interventions/improvements to reduce the gap in the population. Overall, results demonstrated that some segments of the population - foremost poorest Southern rural populations - still have limited access to water and are forced to practise open defecation. Finally, we suggest to include standardized indicators that address safely managed drinking water and sanitation services in future surveys and studies to increase accuracy of national estimates.

## 1. Introduction

Access to safe water and basic sanitation is indispensable for human life and dignity and has been recognized as such through the Human Right to Water and Sanitation (UN Committee on Economic Social and Cultural Rights 2010). Efforts to improve access were accelerated as a result of with the Millennium Development Goals (MDGs) world commitments. Yet, lack of access to safe drinking water and adequate sanitation remain widespread global issues. According to the WHO and the UNICEF Joint Monitoring Programme (JMP), in 2015, 663 million people were estimated to live without access to an improved water source and 2.4 billion people were still lacking access to improved sanitation facilities. Target 7C of the MDGs was to halve, by 2015, the proportion of the population without sustainable access to safe drinking water and basic sanitation (i.e. improved water supply and sanitation services). Despite the MDG Target 7C achievement with regard to drinking water, 42.5% of the world population did not have access to water on premises and needed to fetch it (UNICEF/WHO 2015). For sanitation, the target was not achieved although significant improvements were observed. Open defecation (OD) continued to be practised by 13% of worldwide population among which nine out of ten were living in rural areas (UNICEF/WHO 2015).

The apparent global progress in achieving the MDGs relative to drinking water masks the very limited progress in some segments of the population. Rapid urbanization worldwide has led to the necessity of improving urban services but has also exacerbated inequalities between urban and rural populations (Bain et al. 2014b, Wolf et al. 2013). At the end of the MDGs, further progress was called for through the inauguration of the Sustainable Development Goals (SDGs) that will extend until 2030. The latter’s targets aim to: achieve equitable universal access to safe and affordable drinking water; achieve access to adequate and equitable sanitation and hygiene for all and end open defecation, paying special attention to the needs of women and girls and those in vulnerable situations by 2030.

The population using *safely managed drinking water* (i.e., an improved water source which is located on premises, available when needed and free from faecal and priority chemical contamination) and *safely managed sanitation services* (i.e., improved facilities which are not shared with other households and where excreta are safely disposed *in situ* or transported and treated off-site) were set as improved indicators used by JMP to monitor progress towards the SDGs. This adjustment reduced the baseline proportion of the population considered as having access to safe and affordable drinking water (71% of the global population) or adequate and equitable sanitation (39% of the global population) in comparison to the MDGs, but is likely to increase the accuracy of the indicators (WHO/UNICEF 2017). The current population with access to safely managed drinking water can be further broken down to 85% and 55% in urban and rural areas. With regard to safely managed sanitation service, the proportion of the global urban and rural population with access was determined to be 43% and 35%, respectively (WHO/UNICEF 2017).

Agenda 2030 called for “leaving no one behind” (WHO/UNICEF 2017), advocating for efforts aimed at reducing the service gaps and reaching the populations who are furthest behind (i.e., low income, rural, etc.). Households using unimproved drinking water sources including surface water and practising open defecation are the most likely to need assistance. As primarily responsible for households’ water supply, sanitation and hygiene, women and girls’ lives are even further driven by a lack of access to basic services (Chipeta 2009, Graham et al. 2016).

Access being usually greater in urban areas, global trends show that the service level of water and sanitation increases with the wealth of populations (Cassivi et al. 2018b, Oageng and Mmopelwa 2014, Seyoum and Graham 2016, WHO/UNICEF 2017). Factors such as climate change and population growth are expected to increase vulnerability to water stress and affect the adaptability of sanitation systems, especially for vulnerable populations in low- and middle-income countries and other low resource contexts with limited coping strategies (Fulco 2009, Sherpa et al. 2014).

Malawi is ranked among the poorest countries (GDP per capita = 389 US$) and relies greatly on official development assistance (World Bank 2018); it has experienced a significant population growth in the last decades. Water supply and sanitation facilities have improved following major investments in the sector, however, foreign aid interventions and allocations in water supply and sanitation services weren’t provided to the areas with the highest needs as most were implemented in settings with greater existing infrastructure (Marty et al. 2017, Wayland 2017). The MDG water-related target was achieved whereas moderate progress was made in sanitation (UNICEF/WHO 2015). In 2015, it was reported that 87% of the national population had access to an improved water source (i.e. piped water, boreholes or tubewells, protected dug wells, protected springs, rainwater, and packaged or delivered water) and 41% had access to an improved sanitation facility (i.e. flush/pour flush to piped sewer system, septic tanks or pit latrines; ventilated improved pit latrines, composting toilets or pit latrines with slabs)(WHO/UNICEF 2018). As such, it is likely that these statistics were driven by the progress made in areas that did not have the greatest need.

With regard to the SDGs, the number of people with safely managed drinking water and sanitation services in Malawi is expected to decrease from the old improved/unimproved metric, when accounting for additional criteria of access (Cassivi et al. 2018a, Cassivi et al. 2017, Smiley 2017). Unfortunately, information on safely managed services in the country remains unknown due to data unavailability (e.g. water quality and contamination, excreta disposal and treatment). Significant inequalities are nevertheless expected to rise as in similar contexts where water quality is generally worse in rural areas (Bain et al. 2014a, Bain et al. 2012, Bain et al. 2014b). Furthermore, the use of improved water and sanitation facilities is greater in the wealthiest quintiles, confirming the necessity to target the most vulnerable populations (WHO/UNICEF 2017).

There is a need to focus on the variation in lack of access to water and open defecation in the different geographic and socio-economic groups of the population using classification such as region, development type (urban/rural), wealth or education level (Galan et al. 2013, Hopewell and Graham 2014). However, research focusing on disaggregated data is limited, and the extent of such variations is not well known. Using the case study of Malawi, we compare the indicators of access to water and sanitation across those four categories. Further, as individuals who have neither good water access nor good sanitation are those who should be first addressed, we consider these two measures to identify where priority should be placed for interventions/improvements.

The objectives of this study are to describe emerging trends on progress and inequalities in water supply and sanitation services over a 25-year period (1992 - 2017) and to identify areas where proportion of the population without basic access to water and practising open defecation remains the most vulnerable in Malawi. This is a timely exercise to identify and target groups that were left behind and help reach, by 2030, the targets set in the SDGs.

### 2. Methods

Data sources used for this study were the UNICEF Multiple Indicator Cluster Surveys (MICS), USAID Demographic and Health Surveys (DHS), and Malaria Indicators Survey (DHS-MIS), all of which are harmonized household surveys. Household data for Malawi were downloaded and extracted for each year publicly available (Table 1) (UNICEF 2019, USAID 2019). The use of longitudinal data required pre-processing as MICS and DHS questionnaires have been modified and conducted at different time of year. Data from each year were systematically recoded and categorized to ensure homogeneity of the variables prior to appending. The JMP methodology for compilation and classification was replicated to produce the most accurate estimates (WHO/UNICEF 2018). All datasets were cleaned and appended for harmonization and descriptive and analytical statistics were conducted in StataSE 14.

**Table 1.**
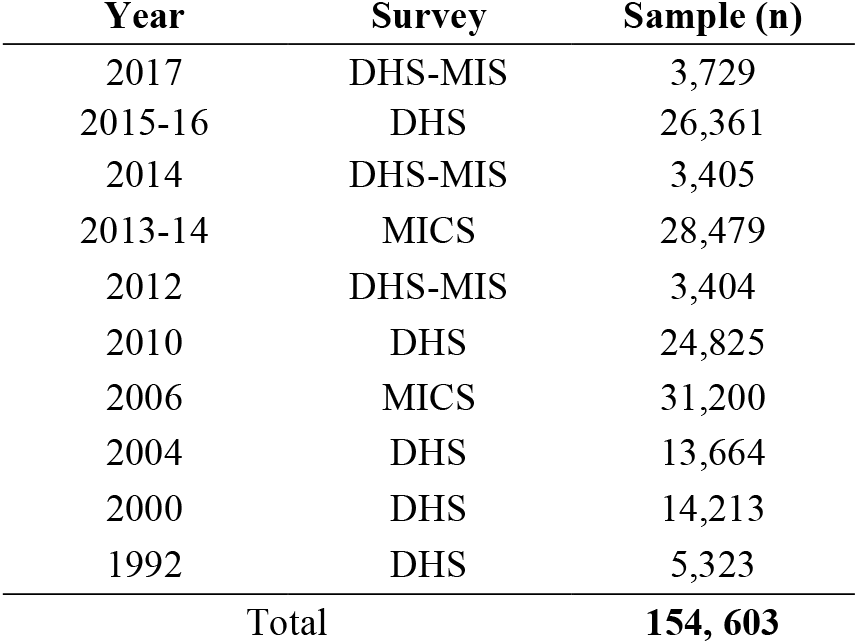
Information on data sources included in analysis.

| **Year** | **Survey** | **Sample (n)** |
| --- | --- | --- |
| 2017 | DHS-MIS | 3,729 |
| 2015-16 | DHS | 26,361 |
| 2014 | DHS-MIS | 3,405 |
| 2013-14 | MICS | 28,479 |
| 2012 | DHS-MIS | 3,404 |
| 2010 | DHS | 24,825 |
| 2006 | MICS | 31,200 |
| 2004 | DHS | 13,664 |
| 2000 | DHS | 14,213 |
| 1992 | DHS | 5,323 |
| **Total** |  | **154, 603** |

Relevant data on drinking water supply and sanitation facilities were disaggregated to target the most vulnerable populations and emphasize the lingering lack of access (Eagin and Graham 2014). The principal indicators were reduced to the following due to their availability and consistency across surveys over time: 1) the proportion of the population without access to an improved water source located within 30 minutes (i.e., basic access); and 2) the proportion of the population practising open defecation. Appended datasets included a total sample of 154,603 Malawian households covering a period of 25 years.

DHS and MICS surveys are designed with a stratified, two-stage cluster design, which aims to be representative of geographic regions, urban/rural areas and wealth quintiles. Household weight as reported by DHS and MICS was multiply by the number of household members to calculate population weight (WHO/UNICEF 2018). Population weighting was used to produce estimates of coverage for access to water and open defecation at the individual level, as applied to monitor global progress in access to drinking water and sanitation services.

Trends in access to water and open defecation were assessed using explanatory variables available in DHS and MICS surveys. Analysis was conducted by grouping data in regions, areas (i.e., urban/rural), wealth index (i.e. composite measure of a household’s cumulative living standard as defined by DHS and MICS) and education levels (i.e. no education, primary education, secondary education and higher). According to the National Statistical Office urban areas are Lilongwe City, Blantyre City, Mzuzu City, and the Municipality of Zomba. Analysis was limited to variables that were available across the data sets. Variables used to conduct analysis were: source of drinking water (*n*=152 160), time to get to the water source (minutes) (*n*=150 600) and type of toilet facility (*n*=152 097). Classification of improved and unimproved water technologies followed JMP classification for SDG monitoring. Therefore, ambiguous categories that were used in the first rounds of surveys (i.e., 1992, 2000, 2004) were classified as follows: “spring” and “public well” as unimproved water technologies and “traditional pit latrines” as unimproved sanitation technology.

## 3. Results

### 3.1. Trends and Progress

#### 3.1.1. Drinking Water

The proportion of the population without basic access to drinking water (i.e., improved water sources within 30 minutes) decreased from 1992 to 2017 reflecting considerable progress (Table 2). At the national level, the population without access to an improved water source within 30 minutes walking distance decreased from 57% to 22% representing a percentage change of 62.01%. The greatest improvement was observed between 2015 and 2017 where the proportion of the population without access considerably decreased at the national level reflecting similar change at the disaggregated level. Although household access was improved, the proportion of women who were responsible for fetching water remained roughly the same over time (varying between 84 and 87% from 2006 to 2014).

**Table 2.**
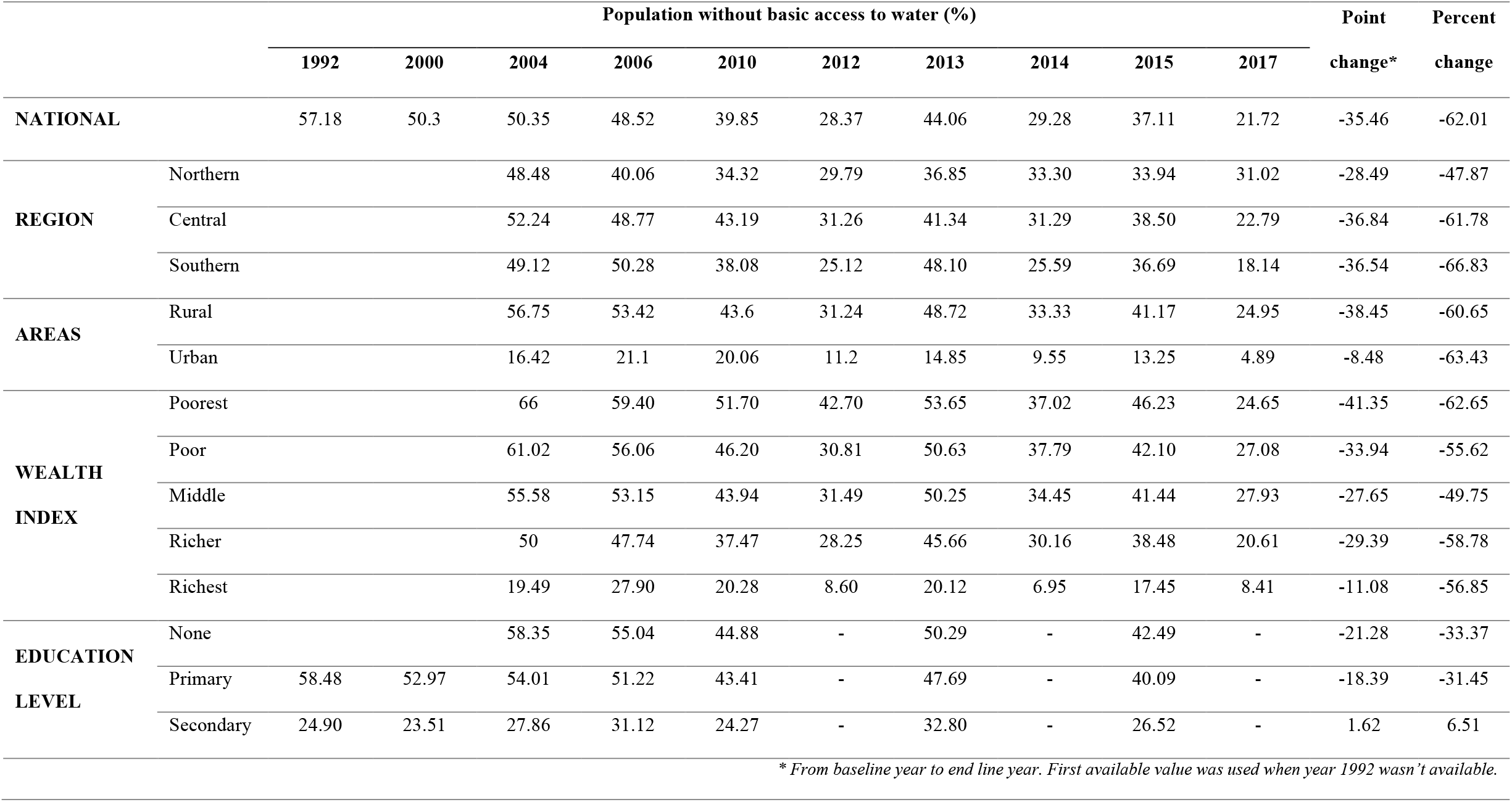
Population without basic access to drinking water (1992-2017) (DHS, MICS)

Progress towards providing basic access to water was greater in rural areas– where 83% of the population lives (World Bank 2018) – than in urban areas with a decrease of 38% bringing the proportion of the population without access down to 25% in 2017. Although the percentage point change from 1992 to 2017 was greater in rural areas, the percentage change attributable to improving access in urban areas was higher with a decrease from 13% to 5%. The gap between urban and rural populations remains one of the greatest among all groups although relative percent change was similar (40% relative change) over time.

Across regions, similar progress was made in the Central and Southern regions – where respectively 43% and 44% of the population lives– as the proportion of the population without basic access was reduced by 37% (NSO Malawi 2008). Less progress was observed in the Northern Region where the population without basic access decreased from 60% to 31%. The proportion of the population without access remains the highest in this region accounting for the smallest population in the country.

The most important progress within the wealth quintiles was among the poorest group in which the percentage of the population without access was lower than in poor and middle upper wealth quintiles. At the opposite end, the richest group had the lowest progress in terms of percentage point change, which is explained by already high levels of service. Overall, the proportion of the population without access to an improved water source within 30 minutes was at least halved in all groups.

Significant disparities were observed between education levels: the proportion of the population without basic access increased between 1992 and 2017 in households with secondary or higher level of education. This is likely explained on account of the fact that the latter are probably urban and unemployed while most programming targets rural uneducated. Progress was, however, observed in groups without education and primary education. Although the percentage of the population without basic access was reduced by one third from 1992 to 2015 for all groups, it remained higher than for the population with secondary level education. The difference in access to drinking water with regard to education illustrates the highest inequalities among all groups.

#### 3.1.2. Open Defecation

The number of people practising open defecation (OD) was reduced at the national scale from 1992 to 2017 (Table 3). Over the 25-year period, OD was reduced by 19% (a reduction from 25% to 6%). An important difference between rural and urban areas was observed in 1992, where 28% and 2% of the population were practising open defecation, respectively; this gap then narrowed in the following years. It is possible that efforts were made towards improving access to sanitation in rural areas where most of the population lives. The percentage of the population practising open defecation was reduced by three quarters, dropping from 28% to 7% from 1992 to 2017. Although the proportion of people practicing OD was already low in urban areas, it was nearly halved over the 25-year period reaching 1% of the population.

**Table 3.** Population practising open defecation (1992-2017) (DHS, MICS)

|  |  | Population practising open defecation (%) |  |  |  |  |  |  |  |  |  | Point | Percent |
| --- | --- | --- | --- | --- | --- | --- | --- | --- | --- | --- | --- | --- | --- |
|  |  | 1992 | 2000 | 2004 | 2006 | 2010 | 2012 | 2013 | 2014 | 2015 | 2017 | change* | change |
| **NATIONAL** |  | 24.76 | 16.41 | 14.45 | 12.08 | 9.83 | 13.45 | 4.90 | 11.27 | 5.33 | 6.02 | -18.74 | -75.69 |
| **REGION** | Northern | 18.41 | 13.41 | 14.29 | 11.79 | 13.21 | 14.29 | 11.79 | 13.21 | 14.01 | 3.64 | -14.77 | -80.23 |
|  | Central | 28.19 | 17.89 | 16.25 | 10.75 | 10.69 | 13.02 | 4.30 | 10.25 | 4.34 | 4.86 | -23.33 | -82.76 |
|  | Southern | 23.59 | 15.81 | 12.79 | 13.45 | 8.10 | 13.70 | 5.70 | 9.31 | 6.23 | 7.66 | -15.93 | -67.53 |
| **AREAS** | Rural | 27.97 | 18.88 | 16.22 | 13.83 | 11.23 | 15.12 | 5.60 | 13.03 | 6.15 | 6.94 | -21.03 | -75.19 |
|  | Urban | 2.23 | 1.44 | 5.05 | 2.26 | 2.46 | 3.44 | 0.49 | 2.69 | 0.50 | 1.24 | -0.99 | -44.39 |
| **WEALTH INDEX** | Poorest | - | - | 42.78 | 25.22 | 26.18 | 31.45 | 15.13 | 22.51 | 15.30 | 15.09 | -27.69 | -64.73 |
|  | Poor | - | - | 15.99 | 16.48 | 11.60 | 17.05 | 4.76 | 16.63 | 6.03 | 5.16 | -10.83 | -67.73 |
|  | Middle | - | - | 7.68 | 10.68 | 7.30 | 11.20 | 3.17 | 11.67 | 2.80 | 7.43 | -0.25 | -3.26 |
|  | Richer | - | - | 5.16 | 6.72 | 3.74 | 6.79 | 1.32 | 5.23 | 2.24 | 1.78 | -3.38 | -65.50 |
|  | Richest | - | - | 0.81 | 1.7 | 0.6 | 0.84 | 0.1 | 0.33 | 0.35 | 0.65 | -0.16 | -19.75 |
| **EDUCATION LEVEL** | None | 35.59 | 26.88 | 21.33 | 18.17 | 14.92 | - | 8.88 | - | 8.38 | - | -27.21 | -76.45 |
|  | Primary | 22.40 | 15.16 | 15.02 | 12.22 | 10.44 | - | 5.22 | - | 5.80 | - | -16.6 | -74.11 |
|  | Secondary | 3.56 | 3.10 | 3.58 | 4.03 | 3.15 | - | 1.59 | - | 2.19 | - | -1.37 | -38.48 |

\* From baseline year to end line year. First available value was used when year 1992 wasn't available.

Progress was greater in the central region where OD prevalence was the highest at the baseline year with a percentage change of 83% (between1992-2017). Lower progress was observed in the Southern Region which has highest proportion of the population practising open defecation.

With regard to the wealth index quintiles, the proportion of the population practising open defecation was reduced by more than three quarters in all groups except the middle wealth group where the proportion was nearly halved. The gap between the poorest and richest quintile was reduced between 1992 and 2017. The most important percentage change was observed in the richest group reaching nearly 0% in 2017.

Open defecation was reduced by three quarters in the population without education or with primary education reduced to fewer than 10% in 2015. Progress cover the 25-year period was lower in households where in the household head had secondary or higher education.

#### 3.1.3. Water and Sanitation

The trends show that the proportion of the population without access to an improved water source within 30 minutes and practising open defecation was even further reduced than individual progress in water and sanitation areas (Table 4). At the national scale, the percentage of the population without basic access to water and practising open defecation decreased nearly 90% from 16% to 2% over the 25-year period. Results show important progress in demonstrating improvement in access for either or both water and sanitation among the populations that were further behind.

**Table 4.** Population without basic access to water and practising open defecation (1992-2017) (DHS, MICS)

|  |  | Population without basic access to water and practising open defecation (%) |  |  |  |  |  |  |  |  |  | Point | Percent |
| --- | --- | --- | --- | --- | --- | --- | --- | --- | --- | --- | --- | --- | --- |
|  |  | 1992 | 2000 | 2004 | 2006 | 2010 | 2012 | 2013 | 2014 | 2015 | 2017 | change* | change |
| **NATIONAL** |  | 16.07 | 10.12 | 9.06 | 7.41 | 4.64 | 4.49 | 2.76 | 4.73 | 2.45 | 1.75 | -14.32 | -89.11 |
| **REGION** | Northern | 13.19 | 8.96 | 8.25 | 5.76 | 4.41 | 4.28 | 2.03 | 9.44 | 2 | 1.81 | -11.38 | -86.28 |
|  | Central | 19.63 | 11.43 | 10.22 | 6.43 | 5.27 | 5.37 | 2.17 | 3.82 | 2.06 | 1.53 | -18.1 | -92.21 |
|  | Southern | 13.94 | 9.22 | 8.20 | 8.76 | 4.08 | 3.71 | 4.42 | 3.49 | 2.92 | 1.94 | -12 | -86.08 |
| **AREAS** | Rural | 18.23 | 11.68 | 10.36 | 8.54 | 5.35 | 5.11 | 3.17 | 5.48 | 2.85 | 2.08 | -16.15 | -88.59 |
|  | Urban | 0.87 | 0.65 | 2.16 | 1.08 | 0.88 | 0.82 | 0.19 | 1.11 | 0.1 | 0.03 | -0.84 | -96.55 |
| **WEALTH INDEX** | Poorest | - | - | 28.40 | 15.54 | 13.55 | 13.65 | 8.69 | 10.02 | 7.58 | 4.52 | -23.88 | -84.08 |
|  | Poor | - | - | 9.88 | 10.42 | 5.03 | 5.10 | 2.52 | 6.80 | 2.51 | 1.66 | -8.22 | -83.20 |
|  | Middle | - | - | 4.03 | 6.13 | 2.99 | 2.05 | 1.86 | 5.40 | 1.23 | 2.13 | -1.9 | -47.15 |
|  | Richer | - | - | 2.65 | 4.33 | 1.59 | 1.70 | 0.68 | 1.44 | 0.88 | 0.43 | -2.22 | -83.77 |
|  | Richest | - | - | 0.45 | 0.89 | 0.16 | 0 | 0.05 | 0 | 0.09 | 0.03 | -0.42 | -93.33 |
| **EDUCATION LEVEL** | None | 22.86 | 17.14 | 14.14 | 11.38 | 7.25 | - | 4.81 | - | 4.23 | - | -18.63 | -81.50 |
|  | Primary | 14.72 | 9.08 | 9.34 | 7.40 | 4.93 | - | 3.05 | - | 2.64 | - | -12.08 | -82.07 |
|  | Secondary | 2.01 | 1.92 | 1.48 | 2.36 | 1.24 | - | 0.69 | - | 0.81 | - | -1.2 | -59.70 |

\* From baseline year to end line year. First available value was used when year 1992 wasn't available.

The proportion of the population without access to water and practicing open defecation and progress was similar in all regions over the years. Improvement in the rural areas led to a reduced gap of access between both areas although disparities remain. Most people without access to basic water and sanitation services were living in rural areas below the middle income wealth quintiles.

As education level increased, the proportion of the population without access to water and practicing open defecation was reduced. Progress from 1992 to 2017 was similar in households without education and those with primary education as opposed to secondary level and higher where most of the population already had higher levels of access to water and sanitation at the baseline year.

### 3.2. Population

Although improvements in terms of access to water and open defecation were generally observed between 1992 and 2017, important variations were noted between 2010 and 2017. The proportion of the population without access to an improved water source within 30 minutes and practicing open defecation followed a decreasing trend from 1992 to 2010 and peak fluctuated until 2017 (Figure 1). Further analysis was conducted to investigate the impact of the type of survey data on trends and progress. The observed peaks in data are likely attributable to the type of surveys as years 2012, 2014 and 2017 were conducted under DHS-MIS (Malaria Indicators Survey) with smaller sample sizes. The proportion of the population without access to drinking water reached the lowest values during these years while higher proportions of open defecation were observed. The DHS-MIS survey differed from MICS and Standard DHS by the smaller sample size which may explain fluctuations in estimates. Disaggregation of the DHS and MICS survey data highlights the discrepancy in the distribution of the sample among population groups (Table 5).

**Table 5.** General distribution of the national population drawn for DHS and MICS surveys.

|  |  | General description – National Population |  |  |  |  |  |  |  |  |  |
| --- | --- | --- | --- | --- | --- | --- | --- | --- | --- | --- | --- |
|  |  | 1992 | 2000 | 2004 | 2006 | 2010 | 2012 | 2013 | 2014 | 2015 | 2017 |
| **REGION** | Northern | 28.01 | 16.07 | 11.86 | 19.23 | 17.43 | 15.01 | 18.35 | 28.52 | 18.84 | 33.31 |
|  | Central | 33.14 | 34.01 | 36.25 | 34.62 | 33.75 | 39.48 | 33.46 | 35.3 | 33.21 | 33.33 |
|  | Southern | 38.85 | 49.92 | 51.9 | 46.15 | 48.82 | 45.51 | 48.19 | 36.18 | 47.95 | 33.36 |
| **AREAS** | Rural | 25.15 | 18.86 | 12.62 | 11.18 | 11.72 | 30.99 | 14.31 | 35.57 | 18.93 | 39.96 |
|  | Urban | 74.85 | 81.14 | 87.38 | 88.82 | 88.28 | 69.01 | 85.69 | 64.43 | 81.07 | 60.04 |
| **WEALTH INDEX** | Poorest |  |  | 23.54 | 20.73 | 22.18 | 18.04 | 21.26 | 16.95 | 19.52 | 13.46 |
|  | Poor |  |  | 20.85 | 21.32 | 21.36 | 16.39 | 19.91 | 17.42 | 19.47 | 13.52 |
|  | Middle |  |  | 20.18 | 20.16 | 20.52 | 17.33 | 20.01 | 17.8 | 19.13 | 14.03 |
|  | Richer |  |  | 19.23 | 19.81 | 19.67 | 18.04 | 19.5 | 19.94 | 19.7 | 19.9 |
|  | Richest |  |  | 16.2 | 17.97 | 16.17 | 30.2 | 19.32 | 27.9 | 22.18 | 39.1 |
| **EDUCATION LEVEL** | None | 28.48 | 25.33 | 25.76 | 21.67 | 19.05 |  | 16.41 |  | 15.91 |  |
|  | Primary | 58.64 | 59.46 | 58.04 | 60.12 | 61.03 |  | 61.91 |  | 56.46 |  |
|  | Secondary | 12.87 | 15.2 | 16.2 | 18.21 | 19.92 |  | 21.68 |  | 27.63 |  |

**Figure 1.**
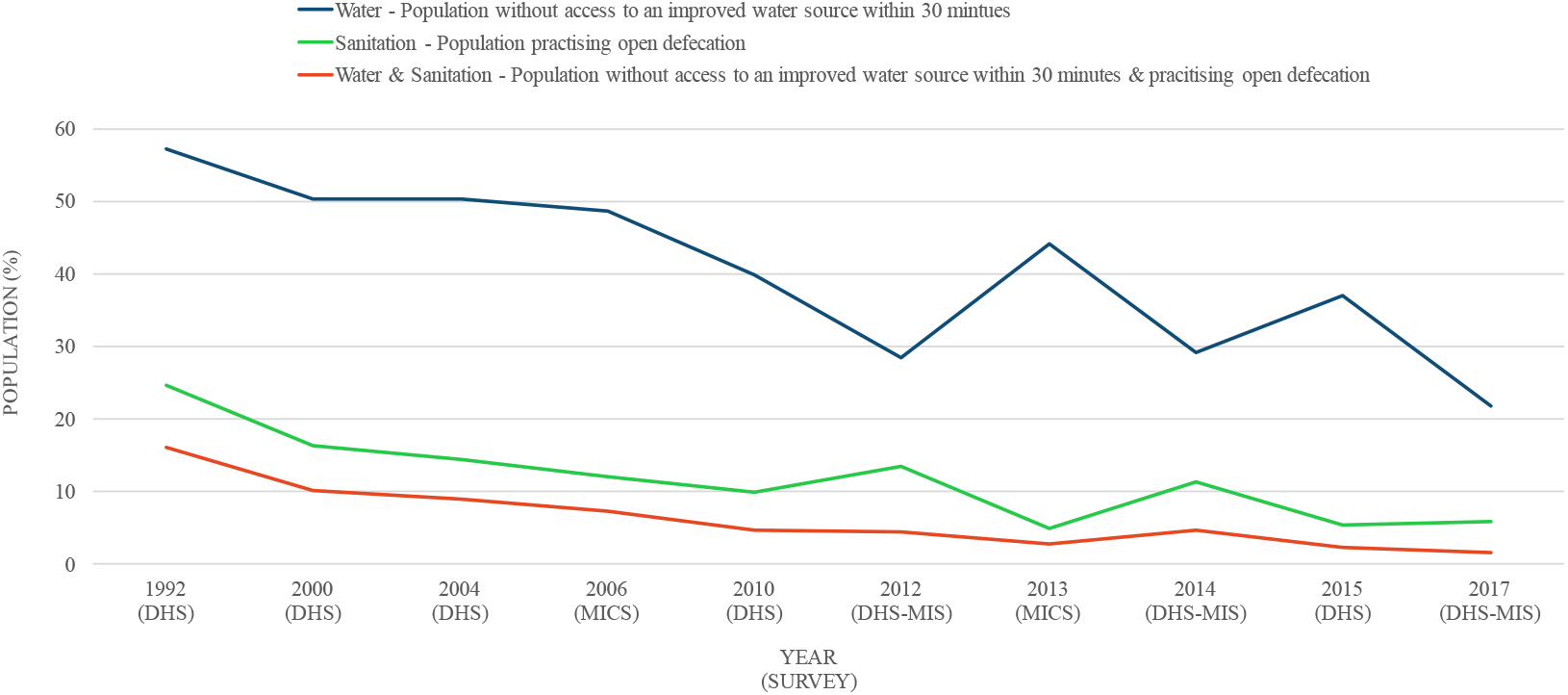
Proportion of the population without access to water and sanitation over the years (1992-2017)

In order to explore how representative the data were, survey data were compared to national population data drawn from the Malawi National Statistics Office Census and The United Nations Population Division’s World Urbanization. A comparison of the total rural/urban population ratio shows an important contrast with survey data particularly in the years where DHS-MIS were conducted (Figure 2). Trends with regards to the rural population in DHS and MICS surveys across time (R^2^=0.1) don’t follow the distribution of the population as reported in national census over the relevant years (1987-2017) (R^2^=0.9). In 2017, the survey population living in rural areas was 60% compared to the actual population where it was estimated to 83%. The rural population in the sample for the years 2012, 2014 and 2017 was underestimated from 15% to 23% compared to census population which likely explains the difference observed in the trends of access to drinking water and sanitation (Figure 2).

**Figure 2.**
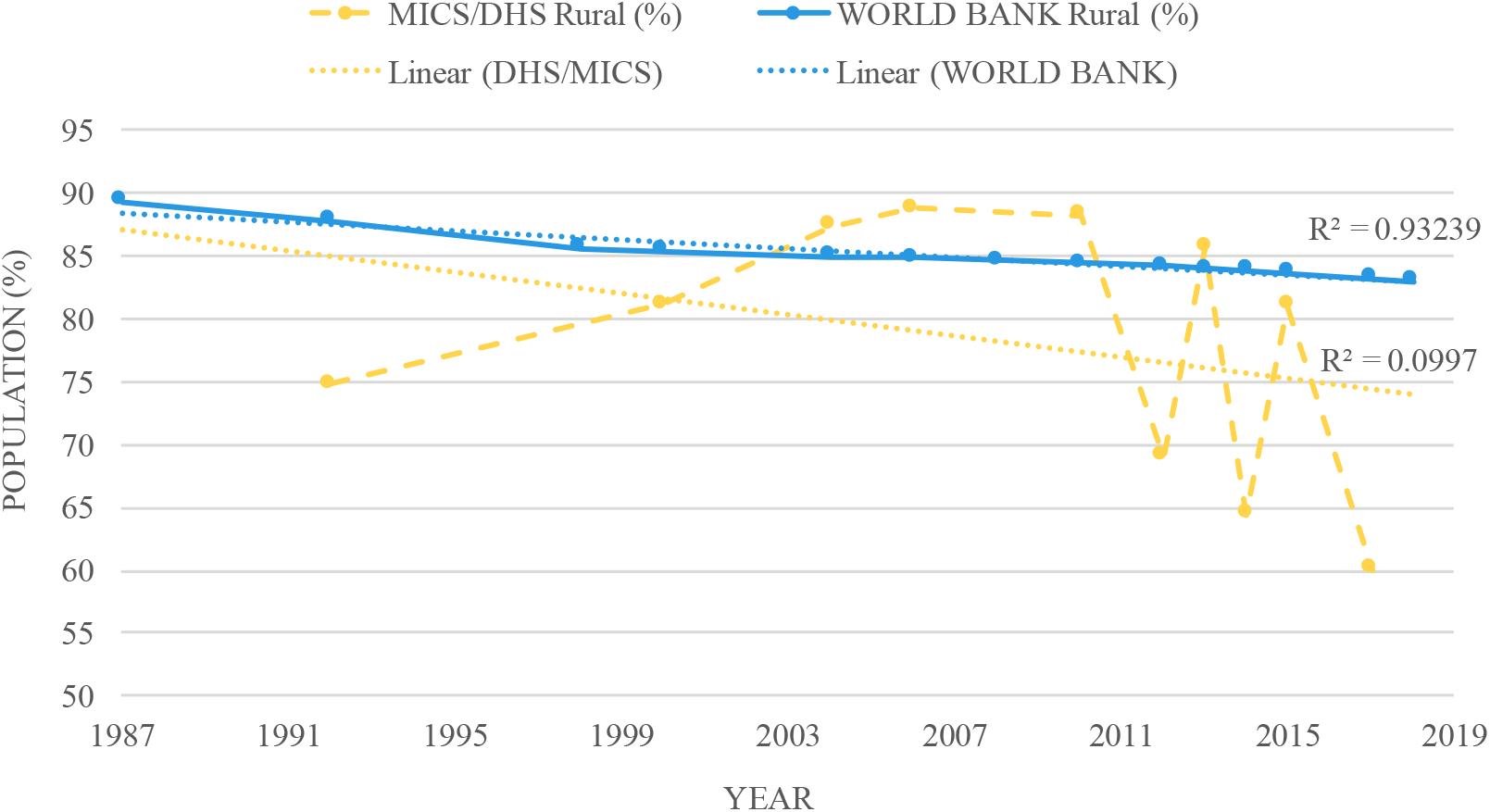
Comparison of the distribution of the rural population in survey sample and total population.

**Figure 3.**
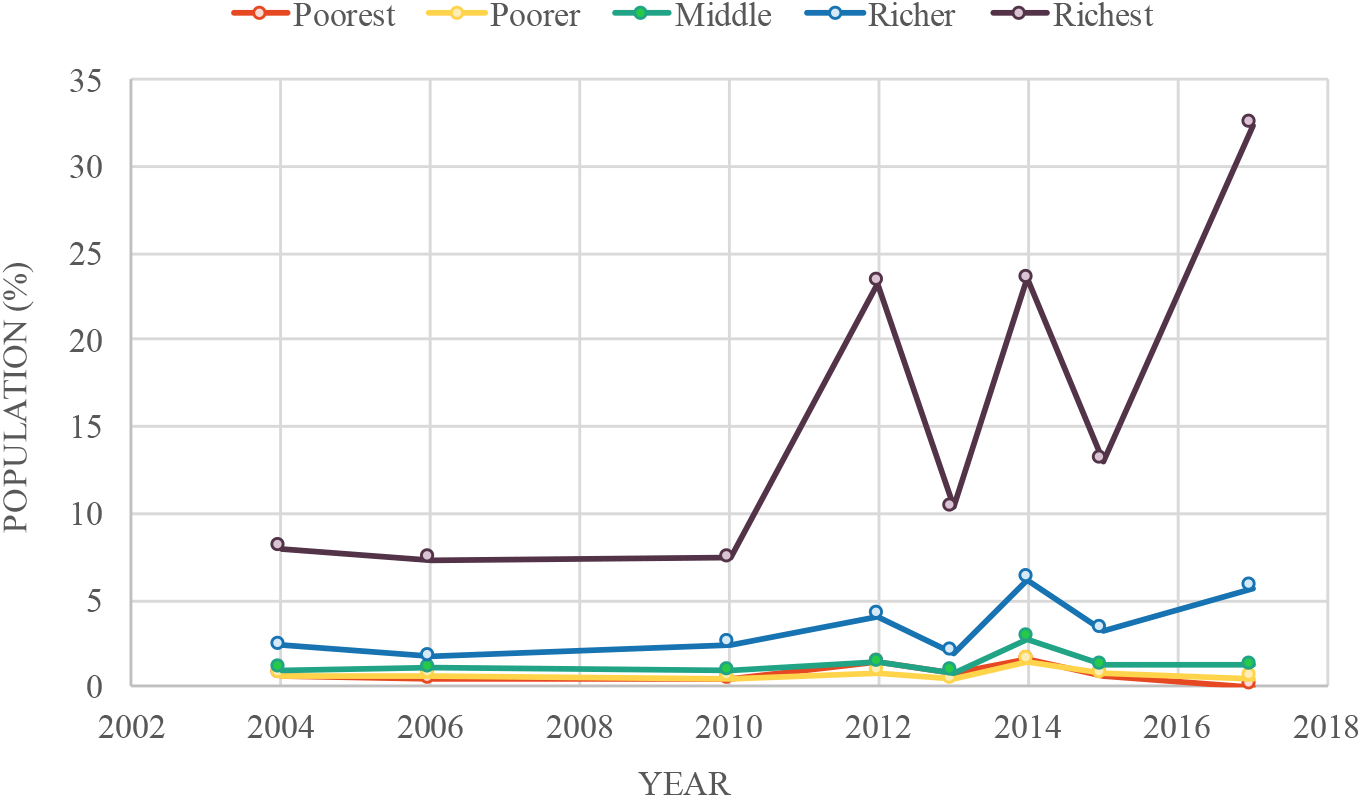
Trends of population by wealth quintiles in urban areas (%)

**Figure 4.**
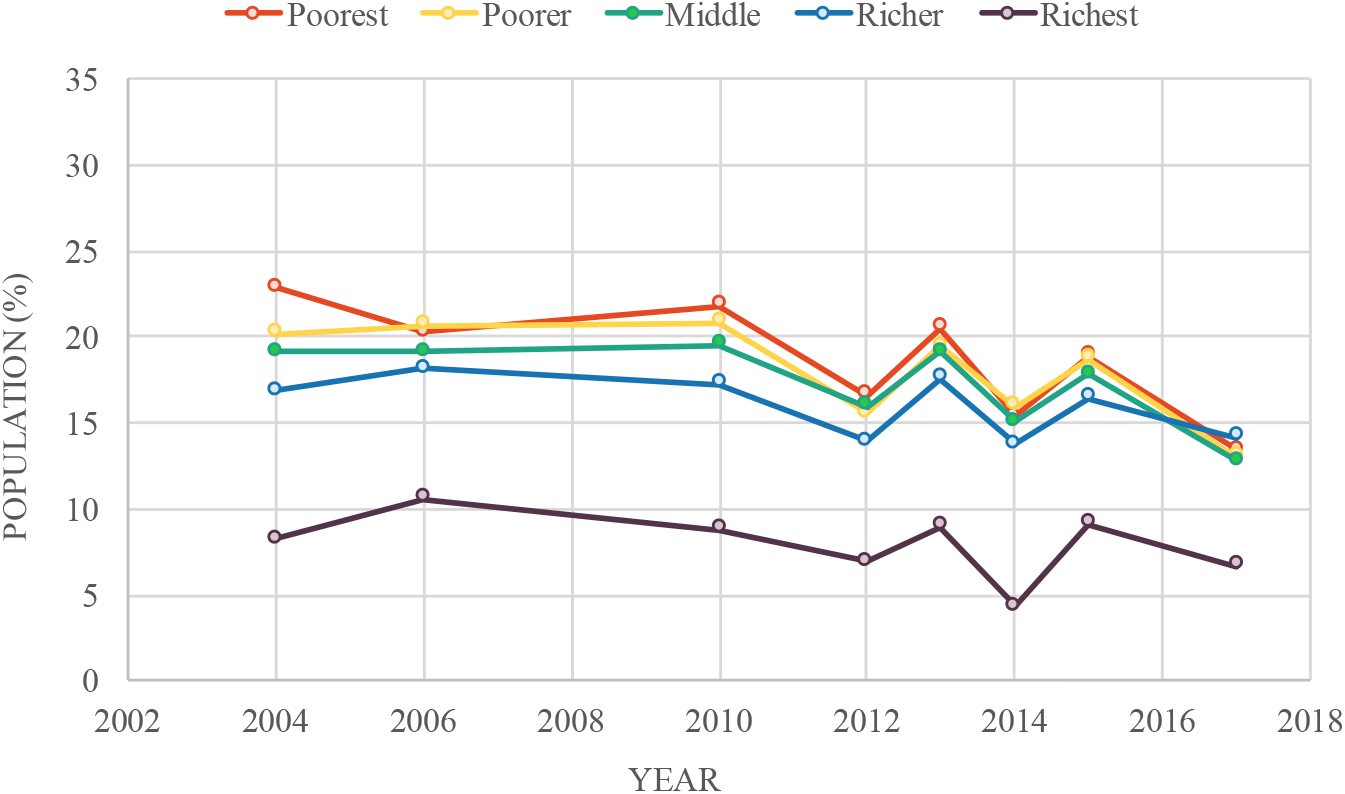
Trends of population by wealth quintiles in rural areas (%)

**Figure 5.**
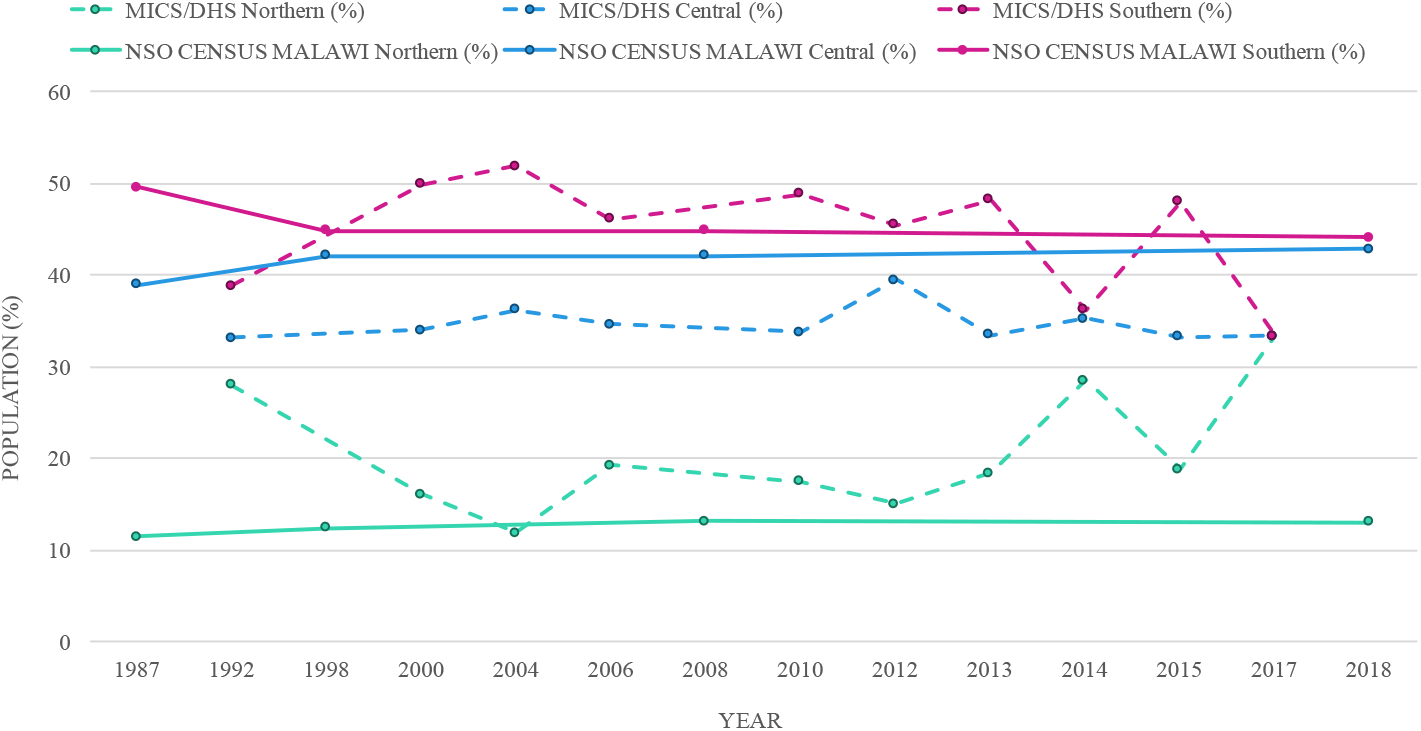
Distribution of the population in the regions with comparison of survey sample and total population.

Variations in the distribution of the population across wealth living standards between rural and urban areas are also observed in the survey sample. These variations are exacerbated in the years where DHS-MIS were conducted. In urban areas, the richest group is overrepresented compared to the other groups which likely represents the actual distribution of the population. The opposite pattern is observed in rural areas where the richest households are underrepresented. The other groups are similar in terms of distribution and generally follow the same trends across the years.

Finally, variations were also observed with respect to the distribution of the sample population in the three regions of Malawi (Table 3). Data from the national population (NSO Census) shows limited changes in the distributions of the population in the different regions of the country over the years. Data from DHS and MICS generally follow the population reported in national census as most inhabitants are located in Central and Southern regions. Differences were, however, observed for the years 1992, 2014 and 2017 where the survey sample was equally distributed among the three regions. Such variations in the sample population don’t represent the population in the country and likely influence the estimates at the country level.

### 3.3. Inequalities and Vulnerability

Following the trend analysis, end-line data were used to further study where lack of access remains the most critical and who is the most vulnerable in the country. Data from DHS 2015 were used as distribution of the sample data was shown to be similar to the national global population than DHS 2017. The largest sample size also demonstrated a higher likelihood of following over last decades’ national estimates.

#### 3.3.1. Drinking Water

Disaggregation of the population without access to an improved water source located within 30 minutes reveals underlying trends and highlights the most vulnerable groups (Figure 6). The population without access is mostly within the Central and Southern regions, with a lower proportion in the Northern region. More than three quarters of the people without access were living in the rural Central or Southern areas. Among all regions, most of the population lacking access lives in rural areas. Interestingly, the populations lacking access were mostly from richer households in the rural, Northern areas. The percentage of the population without access decreased as wealth increased in the Central Region. The population without access to basic drinking was similar in all wealth groups of the Southern Region with the exception of the richest where the proportion was the lowest.

**Figure 6.**
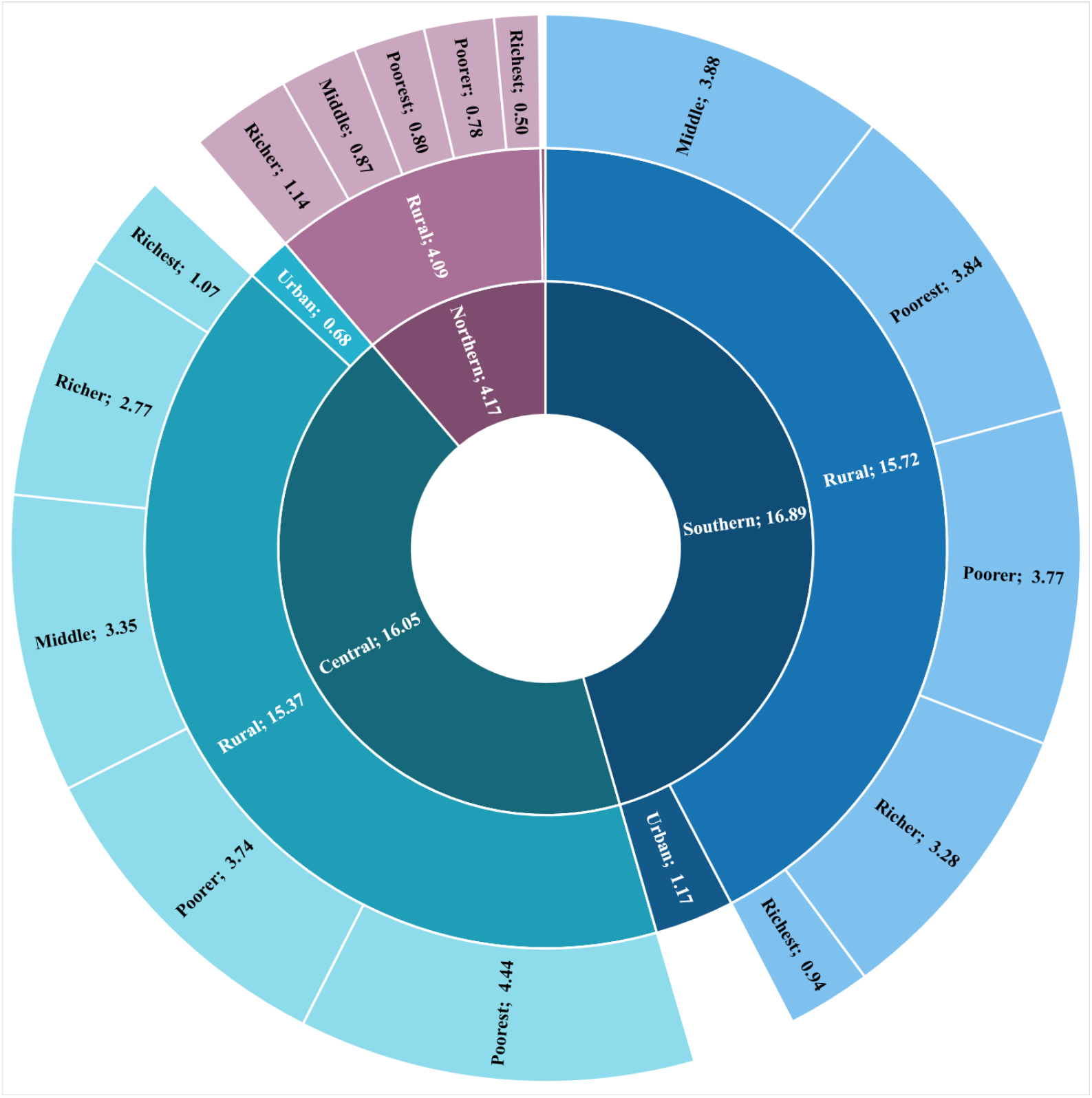
Distribution of population without access to an improved water source located within 30 minutes (%) (2015)

#### 3.3.2. Open defecation

In 2015, 5% of the national population was reported to practice open defecation. Despite significant progress in most of the groups, the distribution of the population practising open defecation was not uniform across groups (Figure 7). Concentrated in the Central and Southern regions, most of the populations practising open defecation were living in rural areas. more than half of the people practising open defecation in Malawi were located in rural Southern areas and nearly one third were living under poorest wealth quintile. There was almost no one in the urban areas and rural richest quintiles practising open defecation in 2015.

**Figure 7.**
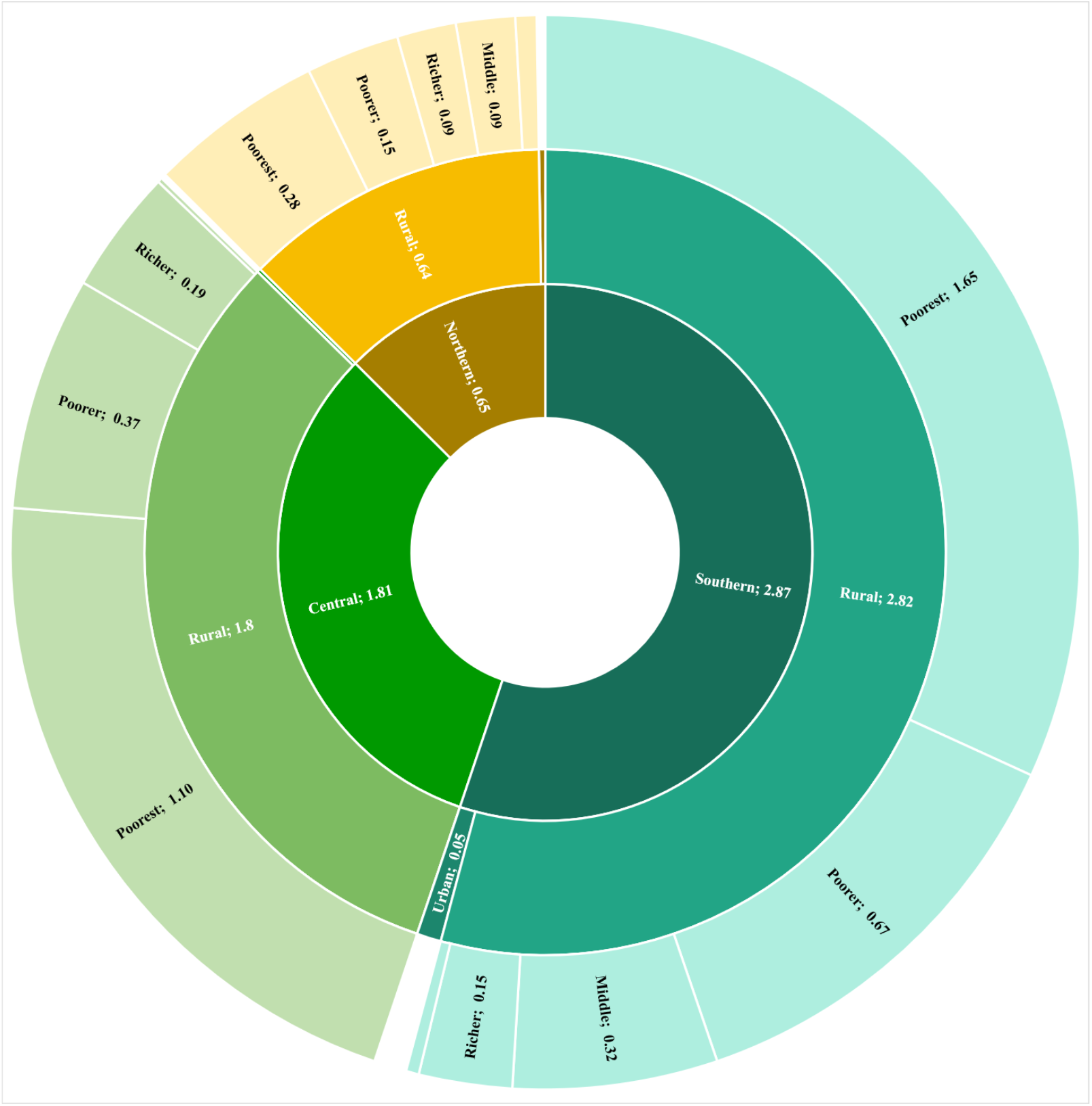
Distribution of the population practising open defecation (%) (2015)

**Figure 8.**
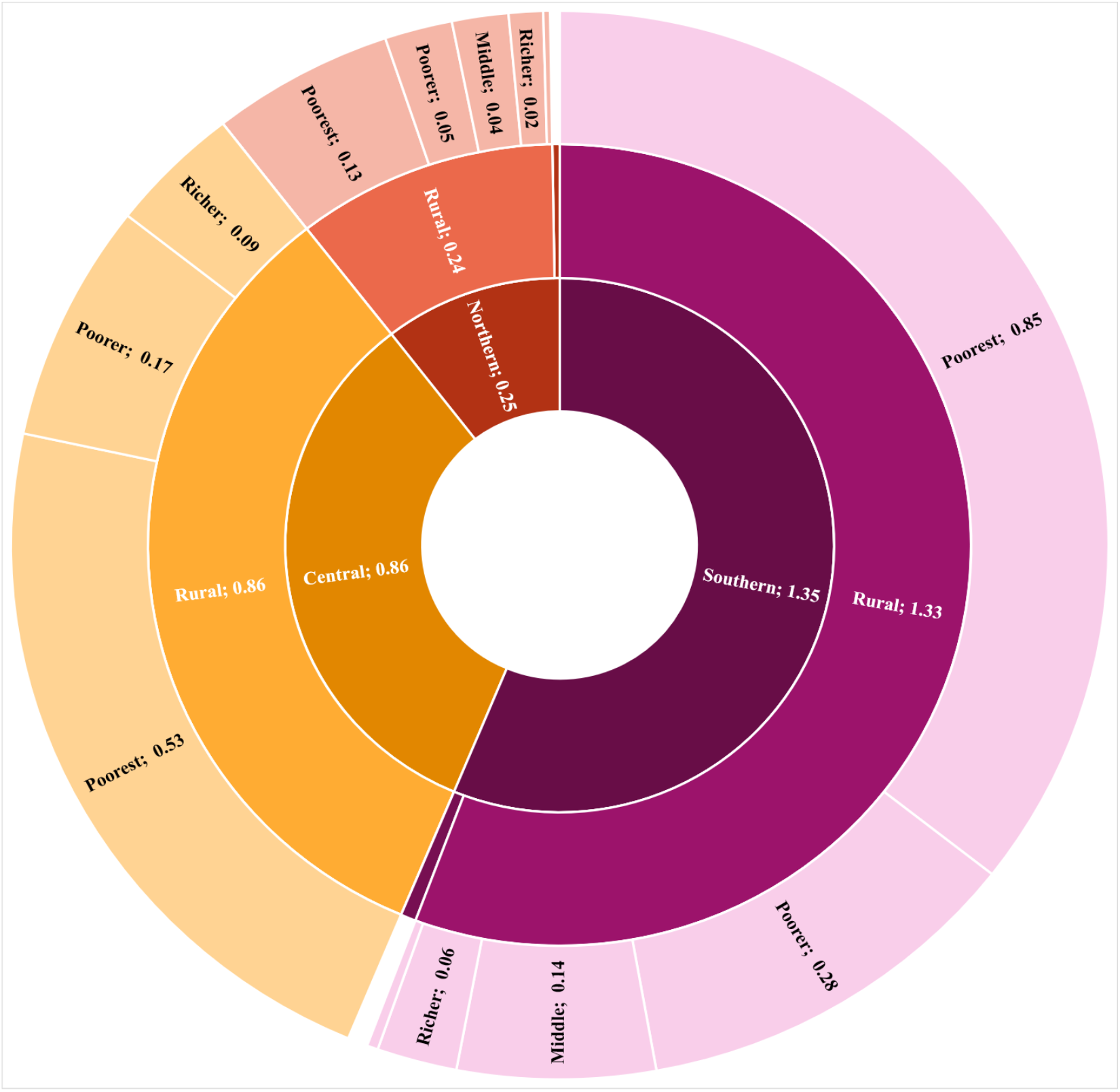
Distribution of the population without access to basic drinking water services and practising open defecation (%) (2015)

#### 3.3.3. Water and sanitation

In 2015, 2% of the population in Malawi didn’t have access to an improved drinking water source within 30 minutes of their household and were still practising open defecation. A general negative association between open defecation and wealth is observed as the practice decreases as wealth increases. More than half of the people living without such access were located in the Southern Region and were mainly part of the poorest quintile of the population. Among all regions, the proportion of the population without access to basic water and sanitation supply was zero (or close to it) in the urban areas and the richer and upper quintiles in all areas.

## 4. Discussion

### 4.1. General Progress

Progress in access to water and sanitation in Malawi is observed in all groups of the population over the 25-year period for which data was available. The largest progress is generally observed in the groups that were furthest behind at the baseline year (e.g., rural, poorest, uneducated), which likely reflects good targeting. Although more people gained access in those groups, i.e. the largest segment of the population, the percentage of the population without access to water or practising open defecation continued to be the highest for those groups, which is consistent with studies conducted in other countries (Eagin and Graham 2014). This reflects that most progress in terms of improving access was observed in the Central and Southern regions. Over time, these regions have narrowed the gap with the Northern Region where lower density along with greater access at the baseline year were reported. Results demonstrated low regional inequalities in terms of access in the country and this is consistent with Pullan et al. (2014) findings.

Improvement in access to basic water and sanitation services in the country is likely attributable to the important investment in the sector over the last decades. Malawi received one of the largest per capita amounts of aid (ODA per capita = 1.07$) within the water sector between 2000 and 2010 (Bain et al. 2013). Galan et al. (2013) demonstrated a significant association between per capita aid disbursement for basic water and sanitation and proportion of the population practising open defecation reduction. The higher proportion of the population lacking access to each water and sanitation separately as opposed to both shows that access was mostly improved for one service – namely water –but not necessarily to each of them. It is evident that improving access to both water and sanitation is indispensable to benefit the population.

The implementation of the Open Defecation Free (ODF) Malawi Strategy 2011 has raised awareness in the country by highlighting the need for involvement from Ministry of Agriculture, Irrigation and Water Development, Ministry of Health, Ministry of Education and key line ministries in collaboration with development partners, NGOs, private sector, communities and key sector players in the sector. Such efforts likely accelerated progress as important change is observed in the rural areas from 2012 onwards. Minimal progress was observed in reducing open defecation in urban areas, and is not surprising considering that ODF Malawi Strategy only focused on rural areas and hasn’t specifically tackled the most vulnerable and marginalized group of the population (e.g. people with disabilities, the elderly, women, children and youth)(Taulo et al. 2018).

Despite the fact that fewer people gained access in groups where access was the greatest at the beginning of the 25-year period, results demonstrated that percentage change improvements still occurred over the years. The constant progress across the different parts of the population, however, demonstrated the positive effect of recent underlying efforts to improve access to water and sanitation in Malawi. The remaining population (though a small percentage of the total) without access to an improved water source within 30 minutes and/or practising open defecation remain the hardest to reach and should be targeted in view of ensuring universal and equitable access for all by 2030.

### 4.2. Urban and Rural Areas

It is evident that the proportion of the population gaining access was larger in rural areas for both water and sanitation services. Different rates of progress in both groups led to a reduction in the gap between rural and urban areas. The population without access to an improved drinking water source within 30 minutes and/or practising open defecation was greatly reduced in both areas. It should be noted that access to water and sanitation was already greater in the urban areas at the beginning of the 25-year period which explains why fewer people gained access. Nevertheless, results demonstrated important progress and similar percentage changes in the urban areas supporting the broad range of people targeted in terms of improvements.

Improvements in access in the urban areas were observed to follow slight urban population growth in Malawi. Overcoming progress despite urbanization may suggest that people moving from rural to urban areas also get at least basic access to water and sanitation or that further efforts were put forward improving access for the vulnerable urban populations. Using global aggregate data, Bain et al. (2013) show that progress in urban household connections may be positively correlated with overall coverage in rural areas.

As improvements may become stagnant with urban population growth, addressing the needs of marginalized populations remains essential to reach universal access (Adams 2018). Previous studies shed a light on the fact that some groups in urban areas may be even more vulnerable than the population in rural areas (Lungu et al. 2019). Lack of access to water and sanitation in urban areas is mainly a burden in peri-urban areas, and this has been documented in Blantyre and Mzuzu (Chipeta 2009, Wanda et al. 2012). The importance of targeting beyond the disadvantaged rural, towards vulnerable urban population has previously been put forward (Chipeta 2009, Lungu et al. 2019) and is reinforced by the data presented here. Further disaggregation of urban populations, by taking into account peri-urban and informal settlements, would strengthen estimates (Bain et al. 2014b) as UN-Habitat (2013) estimated that nearly 70% of the urban population was living in informal settlements in Malawi.

### 4.3. Wealth and Socioeconomic Status

Household wealth and socio-economic status were also observed to influence trends in access to water and sanitation. Generally, a reduction of the population without access to water and sanitation was greater in the lowest quintiles where baseline data was the highest. More people gained access to basic services in the lowest quintiles although the percentage change was similar across quintiles. Results highlight that the different rate changes across quintiles resulted in more equitable access to water across quintiles by narrowing the gap between the poorest and richest populations. Such findings are supported by previous studies that have found a significant association between socio-economic status and access to water and sanitation services (Adams 2018, Eagin and Graham 2014, Seyoum and Graham 2016).

Small variations in the percentage of the population without access to an improved water source were observed but greater inequalities with regard to practising open defecation were highlighted between each wealth quintile. In 2015, the percentage of the population lacking access to improved drinking water sources within 30 minutes in rural areas was almost heterogeneous across quintiles. In the Northern and Southern regions, a greater percentage of the population within the richer and middle quintiles were lacking access to drinking water.

A higher income would generally allow individuals to live where better infrastructure exists. Even where infrastructure for water does not exist, those with greater wealth may still occupy locations with better access to infrastructure such as water supply and sanitation.

The important difference between urban and rural population, however, appears to further highlight inequalities than wealth in the country. These results confirm previous findings which have shown that rural and urban areas faces the strongest inequities (Seyoum and Graham 2016). Understanding the role of socio-economic status will be critical for designing policies targeting the most vulnerable households among the population (Adams 2018). No data are currently available to define the actual distribution of wealth across the country which limits further comparison with the population.

### 4.4. Leave No One Behind

Results from this study demonstrated that although improvements have been made, significant parts of the population still have limited access and are forced to practice open defecation. Such findings highlights the need to target the most vulnerable and marginalized populations (Pullan et al. 2014). Looking at trends by wealth draws attention to the breadth of reconsidering disadvantaged rural thinking (Lungu et al. 2019). Women play a central role in water and sanitation as they continue to perform most of the water fetching. The time and energy associated with fetching water further strengthens gender inequalities and reduces women’s potential for empowerment by limiting opportunities (e.g. education, paid work, healthcare and childcare) and increasing the risk of injuries and exposure to abuse and violence (Curtis 1986, Geere 2015). Women and girls practicing open defecation are also exposed to sexual exploitation and psychosocial stressors, which further compromise their dignity, health and wellbeing (Saleem et al. 2019). Improving access to water, sanitation and hygiene is a key driver to improve women and girls’ lives. Gender equity in terms of access to water and sanitation should be further investigated to address girls and women needs for empowerment. Acknowledging the different context between groups of the population (e.g., urban/rural) and target interventions to appropriate situations is essential to leave no one behind (Adams and Smiley 2018).

### 4.5. Data Limitations

Certain limitations regarding data analysis must be stated. This study used readily available large-scale data collected as part of the MICS and DHS surveys and is thus subject to data quality and accuracy controls. Indicators selected for this study were used as they were the most consistent across the years. Open defecation is not the only one towards safely managed sanitation services and may be considered as the simplest reduction proxy to monitor access to sanitation.

Although the surveys are said to be nationally representative, a variation in data estimates with regards to the type of survey conducted by DHS was observed. The trends of access show a difference between the DHS standard and MIS surveys which is likely attributable to sample size.

It should also be noted that survey estimates are drawn from self-reported data which may introduce biases, such social desirability response bias (e.g. conformity, sensitivity to open defecation, hope for better services), and compromise data reliability and validity (Guest et al. 2005, Van de Mortel 2008). Additionally, the differentiation between areas are based on the country’s urban-rural definition. It remains unclear and important to define whether peri-urban informal settlements are considered as urban or rural areas and perhaps not even included in the sampling. The potential influence of the definition of urban and rural areas may have an impact on estimates especially if the definition was revised over time. It is not possible to compare the trends with the population growth and movement as most of the explanatory variables – regions, area, wealth quintiles - used in this analysis were also used to ensure representativeness of the sample at the study design stage.

## 5. Recommendations

Findings from this study are explanatory and were subject to data availability. Further analysis using additional data sources and more exhaustive indicators and disaggregation factors would be necessary to direct interventions and efforts towards the groups of the population that are still left behind. It is clear that resources should be directed to those who lack minimum access to water and sanitation. In Malawi, such people are mostly found to be in the poorest rural Central and Southern areas.

In view of SDGs, future surveys and studies should include indicators that address safely managed drinking water and sanitation services. A MICS survey along with a water quality module is currently underway in Malawi. The introduction of the first estimates of safely managed drinking water services for the country will likely improve the accuracy of national estimates.

## 6. Conclusions

Progress was made to improve access to drinking water and sanitation in Malawi for all parts of the population as measured by areas, regions, wealth and education. Overall, the proportion of the population without access to an improved drinking water source within 30 minutes and/or practising open defecation was more than halved between 1992 to 2017.

The analysis identified the most vulnerable parts of the population (as measured by having limited access and practicing open defecation) as being poorest rural populations living in Southern Malawi. Emerging trends on progress and inequalities in water supply and sanitation services show that efforts should be put towards improving access to basic services in the most vulnerable populations across all geographic and socio-economic groups of the population. Drawing attention to the people being left behind without at least basic access to water and sanitation service is necessary to reduce the gap within the population and ensure equitable access water and sanitation for all by 2030 with respect to SDG’s agenda.

## 7. Conflict of Interest

Authors have no actual or potential conflict of interest to declare regarding this study.

## Data Availability

MICS and DHS surveys are publicly available online at https://mics.unicef.org and https://dhsprogram.com

## 8. Funding

Authors acknowledge the financial support of the Natural Sciences and Engineering Research Council of Canada (NSERC).

## Notes

### Competing Interest Statement

The authors have declared no competing interest.

